# Alzheimer’s disease CSF biomarkers correlate with early pathology and alterations in neuronal and glial gene expression

**DOI:** 10.1101/2024.06.11.24308706

**Authors:** Ali S. Ropri, Tiffany G. Lam, Vrinda Kalia, Heather M. Buchanan, Anne Marie W. Bartosch, Elliot H. H. Youth, Harrison Xiao, Sophie K. Ross, Anu Jain, Jayanta K. Chakrabarty, Min Suk Kang, Deborah Boyett, Eleonora F. Spinazzi, Gail Iodice, Robert A. McGovern, Lawrence S. Honig, Lewis M. Brown, Gary W. Miller, Guy M. McKhann, Andrew F. Teich

**Affiliations:** Department of Pathology and Cell Biology, Columbia University Irving Medical Center, New York, NY 10032, USA; Taub Institute for Research on Alzheimer’s Disease and the Aging Brain, Columbia University Irving Medical Center, New York, NY 10032, USA; Dept. of Environmental Health Sciences, Mailman School of Public Health, Columbia University, New York, NY 10032, USA; Quantitative Proteomics and Metabolomics Center, Department of Biological Sciences, Columbia University, New York, NY 10027, USA; Department of Neurology, Columbia University, New York, NY 10032, USA; Department of Neurosurgery, Columbia University, New York, NY 10032, USA; Ankyra Therapeutics, Cambridge, MA 02142, USA; Department of Neurosurgery, University of Minnesota, Minneapolis, MN 55455, USA

**Keywords:** Alzheimer’s disease, CSF, biomarkers, proteomics, metabolomics

## Abstract

**INTRODUCTION:** Normal pressure hydrocephalus (NPH) patients undergoing cortical shunting frequently show early AD pathology on cortical biopsy, which is predictive of progression to clinical AD. The objective of this study was to use samples from this cohort to identify CSF biomarkers for AD-related CNS pathophysiologic changes using tissue and fluids with early pathology, free of post-mortem artifact.

**METHODS:** We analyzed Simoa, proteomic, and metabolomic CSF data from 81 patients with previously documented pathologic and transcriptomic changes.

**RESULTS:** AD pathology on biopsy correlates with CSF β-amyloid-40/42, neurofilament light chain (NfL), and phospho-tau-181(p-tau181)/β-amyloid-42, while several gene expression modules correlate with NfL. Proteomic analysis highlights 7 core proteins that correlate with pathology and gene expression changes on biopsy, and metabolomic analysis of CSF identifies disease-relevant groups that correlate with biopsy data..

**DISCUSSION:** As additional biomarkers are added to AD diagnostic panels, our work provides insight into the CNS pathophysiology these markers are tracking.

## 1. Background

Chronic hydrocephalus in the elderly may occur for a variety of reasons, although in the absence of a clear etiology most of these cases are classified as idiopathic normal pressure hydrocephalus (NPH) [1, 2]. Placement of a ventricular CSF shunt can provide symptomatic relief for many elderly patients with NPH [3-5]. At some centers, a cortical brain biopsy is sent to pathology, obtained at the shunt insertion point. Early AD pathology has been reported in a percentage of these biopsies, with β-amyloid plaque pathology ranging from 42% to 67%, and tau pathology relatively sparse in NPH cortical biopsies [6], although some studies have found trace tau pathology at higher levels [7]. Perhaps not surprisingly, AD pathology on biopsy predicts future development of clinical AD, which suggests that at least a subset of the NPH population is in the early stages of AD [8, 9]. For these reasons, AD pathology in NPH patients has been studied by several groups as a way to understand early AD pathophysiology [5, 6, 8-11]. An additional advantage is that biopsy tissue is free of post-mortem changes, and since these are living subjects, longitudinal follow-up after biopsy is possible.

Our group has recently analyzed RNA-seq data from a cohort of 106 NPH patients with varying degrees of accompanying AD pathology [10]. This analysis identified a limited set of genes that correlated with quantified histologic measurements of β-amyloid and tau pathology, with a significant enrichment of immune response genes. Weighted Correlation Network Analysis (WGCNA) identified 4 out of 58 modules that correlated with AD pathology. Two of the modules were enriched for microglial genes, and we found that these two modules partially captured the reported downregulation of homeostatic genes and upregulation of disease-associated microglial (DAM) genes reported in the mouse AD model literature [12]. We also identified an astrocytic module and a neuronal module that correlated with quantified pathology, suggesting that transcriptomic changes accompanying early AD pathology encompass a multi-cellular response with a prominent immune response component (Figure 1). We subsequently validated that these modules correlate with AD pathology in autopsy brain transcriptomic datasets, although our microglial modules capture the downregulation of homeostatic genes reported in the mouse AD model literature better than several autopsy datasets [10]. Our interpretation of these results is that we are capturing an early response to AD pathology in our data, and that this may also be partially captured in the mouse AD model literature. Interestingly, recent snRNA-seq data from human tissue with early pathologic changes has identified a microglial response that is similar to what we reported [11, 13], which further validates the relevance of our data for understanding the early stages of AD.

**Figure 1.**
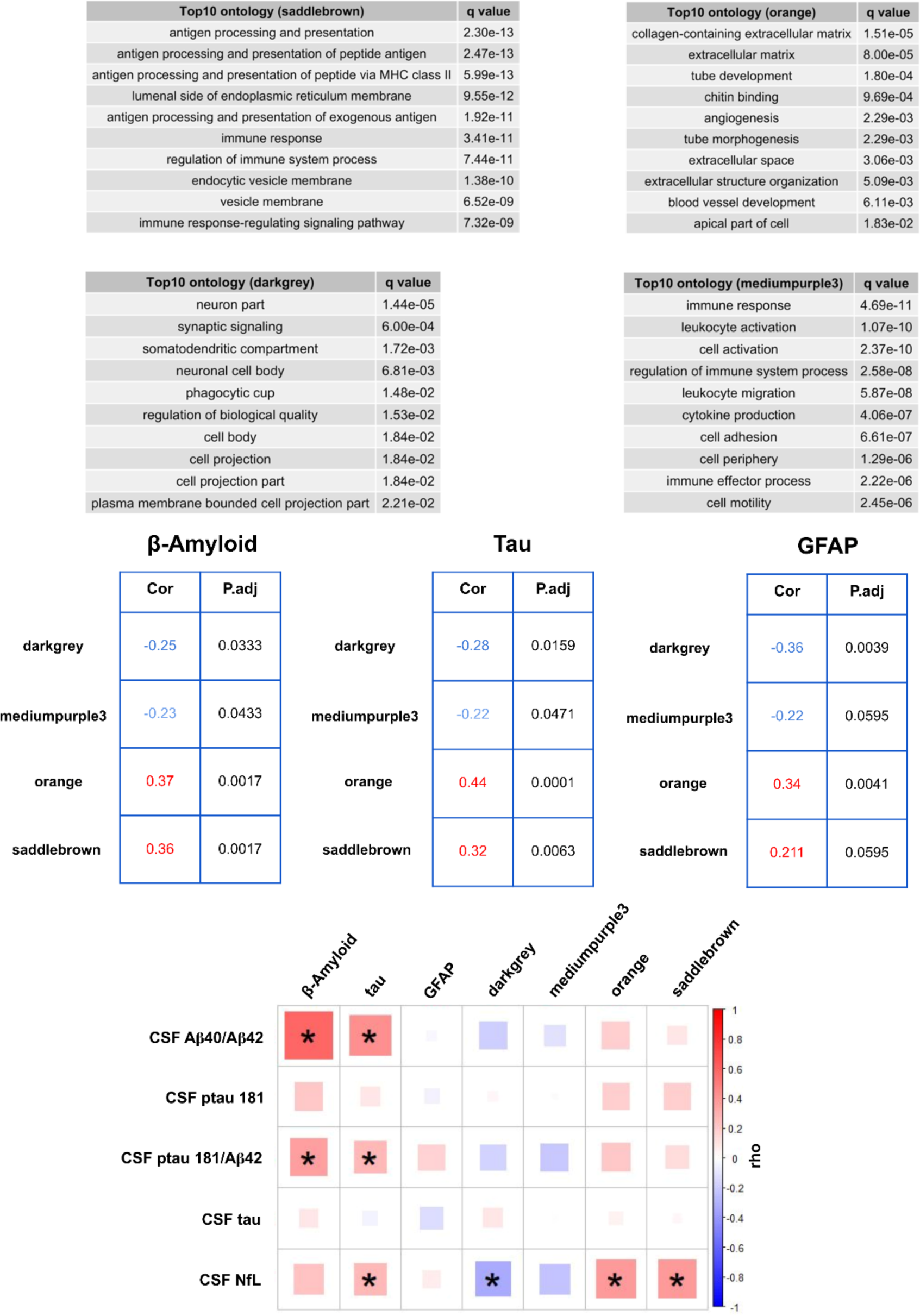
AD pathology, gene expression modules, and AD CSF Simoa markers co-correlate. **A)** Top ontology groups characterizing our four modules reproduced from [10]. Saddlebrown and mediumpurple3 are both enriched for immune response ontology groups, and this is consistent with additional analysis using cell-type specific gene lists that showed that both modules are enriched for microglial genes [10]. We previously showed that the saddlebrown module is enriched for disease associated microglial (DAM) genes identified in AD mouse models, while mediumpurple3 is enriched for homeostatic genes, and this is consistent with saddlebrown positively correlating with AD pathology and mediumpurple3 negatively correlating with AD pathology. The darkgrey module is enriched for neuronal genes, and orange is enriched for astrocytic genes, although this enrichment is less obvious from its ontology analysis. **B)** Our four modules correlate with quantified β-amyloid and tau pathology on the 81 biopsies with CSF similarly to the correlations reported in [10]. For this study, we also added quantified GFAP staining, and correlations with the four modules are shown. In all three cases, FDR adjusted p-values are shown. **C)** Correlations of histologic data and gene expression modules with CSF Simoa measurements of AD biomarkers. * = FDR adjusted p-value < 0.05 (each column adjusted separately). All correlations shown in this figure are Spearman’s rank correlation coefficient. See text for details, and Supplementary Tables 3 and 4 for numbers used in this figure. The n for each Simoa analysis is variable due to some sample failure. In summary, 78 samples have CSF Aβ40/42 values, 80 have CSF ptau 181 values, 77 have CSF ptau 181/Aβ42 values, 80 have CSF tau values, and all 81 have CSF NfL values. GFAP staining was also only achieved on 80 samples. All other analyses here and in the rest of the study are completed on all 81 samples.

In summary, our prior work has produced a useful dataset for understanding early transcriptomic changes in AD, and this has motivated us to identify biomarkers that can track these changes. Our study allows us to ask this question in a unique way, as we are able to obtain CSF at the same timepoint as brain tissue, free of post-mortem artifact. In the present study, we perform proteomics, mass spectrometry-based metabolomics, and Simoa AD biomarker measurements on ventricular CSF that was collected at the same time as the biopsy tissue on 81 NPH patients from our prior study, and analyze this data to identify biomarkers of disease that correlate with biopsy pathology and gene expression changes. We identify a number of previously unreported patterns between brain and CSF, which addresses the challenge of identifying biomarkers for established pathophysiologic CNS changes that occur in the setting of early AD pathology.

## 2. Methods

This study is a retrospective study that uses tissue and CSF samples not required for clinical diagnosis and associated clinical and demographic data. This study was reviewed and approved by the Columbia University Institutional Review Board (IRB), and all relevant ethical regulations have been followed. Processing and analysis of brain tissue, including RNA-seq and histologic analysis of β-amyloid and tau, has been previously reported [10], and is briefly summarized below. Biopsies were taken from frontal cortex in 2/3 of the subjects and parietal cortex in 1/3 of the subjects in our original cohort, with the location for a given patient chosen for cosmetic reasons. This ratio in the subgroup of 81 subjects analyzed in this study is similar. As noted in [10], changes in gene expression that correlate with AD pathology trend similarly in both regions, and we combined all subjects together [10] for purposes of analysis. The average age of our original 106 subjects is 74.9 years, and the subgroup of 81 subjects analyzed in this study (that have both biopsy and ventricular CSF collected) is 74.5 years (see Supplementary Table 1 for patient demographics). CSF analyzed in this study was obtained through the shunt catheter into polypropylene tubes, and promptly frozen and stored at -80°C.

### 2.1 Summary of Previously Reported Data Analyzed in this Study

As reported in [10], RNA was extracted from biopsy samples using miRNeasy Mini Kit (QIAGEN; Cat No./ID: 217004), and samples with RIN values ≥ 6 were selected for sequencing. RNAs were prepared for sequencing using the Illumina TruSeq mRNA library prep kit, and samples underwent single-end sequencing to 30M read depth. The quality of all fastq files was confirmed with FastQC v 0.11.8 [14], followed by variance stabilizing Transformation (VST) [15], and surrogate variable analysis (SVA) [16] and *removeBatchEffect* [17] were sequentially used to remove confounding variables in our dataset. To generate gene expression modules, we utilized <u>W</u>eighted <u>G</u>ene <u>C</u>o-expression <u>N</u>etwork <u>A</u>nalysis (WGCNA) to identify gene co-expression modules [18]. Immunohistochemistry for tau (AT8; Thermo Fisher; Catalog # MN1020), and β-amyloid (6E10; BioLegend; Catalog # 803003) was performed using the Ventana automated slide stainer. β-amyloid plaques were counted per square mm; in slides with enough tissue, three fields were averaged together, whereas in slides with less tissue, the largest number of possible fields were counted. For tau quantification, we devised a rating scale to grade the minimal degree of tau pathology seen in NPH biopsies (Supplementary Figure 1). Grade 0 was given to biopsies with no tau pathology. Grade 1 was given to biopsies that have any tau pathology at all, usually one or more dystrophic neurites, but do not make criteria for Grade 2. Grade 2 was given to biopsies that have at least one tau-positive neuron or neuritic plaque, but do not make criteria for Grade 3. Grade 3 was reserved for biopsies with tau pathology evenly distributed throughout the biopsy.

### 2.2 SIMOA ELISA Analysis

SIMOA technology (Quanterix, Inc., Billerica, MA) on the SR-X platform was used to measure Aβ-40, Aβ-42, and total-tau with the multiplex Neurology 3-plex A kit #101995, NfL with the NF-light Advantage kt (SR-X) kit #103400, and p-tau181 with the p-tau181 Advantage V2 kit #103714. All assays were performed in duplicate for each sample, using 8 calibrators and 2 positive controls (low and high concentrations) in 96-well plates. CSF was rapidly thawed, gently vortexed, centrifuged, and, diluted as per kit specifications depending on assay, and added to kit beads (100 uL) by pipette in each well. Then in succession, plates are incubated for 15 min at 30°C, shaking at 1000 rpm, magnetic-washing 3X for 5 min total, followed by addition of SBG reagent (100 uL), another incubation for 10 min at 30°C at 1000 rpm, washing again 5X for 7 min total and then reading on the SIMOA SR-X machine. Each plate assays in duplicate 34 samples. This highly sensitive assay system has lower limits of quantitation of about 1, 0.1, and 0.1 pg/ml for Aβ-40, Aβ-42, and tau, 0.1 pg/mL for -p-tau181, and 0.3 pg/mL for NfL; coefficients of variation within duplicates are between 3 and 10%. Simoa measurements (Aβ-42/Aβ-40 ratio, total tau, p-tau181, p-tau181/Aβ-42 ratio) were regressed for age and sex with limma::removeBatchEffect (version 3.54.2). Of the 81 CSF samples, a small number of samples failed and we were unable to produce a value. In summary, 78 samples have CSF Aβ40/42 values, 80 have CSF ptau 181 values, 77 have CSF ptau 181/Aβ42 values, 80 have CSF tau values, and all 81 have CSF NfL values.

### 2.3 Proteomics Analysis

Proteins from CSF were studied by quantitative measurement of protein abundance with a mass spectrometry-based proteomic method. Prior to proteome analysis, depletion of high abundance proteins was performed with High-Select Top14 Abundant Protein Depletion Resin columns (Pierce/Thermo Fisher Scientific). CSF proteins were resuspended in 8 M urea, 3 mM dithiothreitol (DTT), 100 mM ammonium bicarbonate in liquid chromatography/mass spectrometry grade water, reduced with dithiothreitol, and alkylated with iodoacetamide. For proteolytic digestion, samples were diluted 5-fold in 100 mM ammonium bicarbonate and then digested using sequencing grade trypsin (Promega V511) at a protease/protein ratio of 1:50 at 37°C for 16 h as described previously [19]. Samples were then desalted with Nest Group C18 Macrospin columns (Southborough, MA). Peptide concentration was evaluated by NanoDrop spectrophotometry (Thermo Fisher Scientific) at 205 nm and LC/MS inject loading amounts were adjusted (normalized) based on peptide concentration. Peptides were separated with an acetonitrile / formic acid gradient at 300 nL / min on a 75 μm ID x 50 cm Acclaim PepMap C18 2 μm particle size column with an UltiMate 3000 RSLCnano liquid chromatograph. This was coupled to a Q-Exactive HF mass spectrometer (Thermo Scientific). Data were acquired in data dependent acquisition mode (DDA) and proteins were identified by database search using PEAKS Studio (version 10.6; Bioinformatics Solutions Inc.) and a Human UniProt reviewed database with isoforms (UniProt release 2020_04, Aug 11 2020). All raw mass spectrometry files produced in this work are publicly available at the MassIVE proteomics repository (https://massive.ucsd.edu).

Protein abundance was measured by label-free quantitation with PEAKS Studio. Peaks software detected 701,261 features across 81 LC/MS/MS runs. Identifications were returned for 1021 proteins with a 1% false discovery rate by the Peaks program. Of the 1021 proteins, 14 proteins subjected to partial antibody depletion, 394 proteins were represented by a single peptide and six added proteins and lab contaminants were deleted from the analysis. 607 total proteins and 528 unique proteins represented by two or more peptides were included in the analysis. No imputation of missing values was performed. Batch correction was achieved using a tunable approach for median polish of ratio (TAMPOR) algorithm [20] for removing technical variance, and protein abundance values were normalized within each batch with no Global Internal Standard (GIS) (1). Effects attributable to age and sex were regressed with limma::removeBatchEffect (version 3.54.2). Log2 normalized protein abundances of 528 CSF proteins from 81 samples were Spearman-correlated to histologic metrics (β-amyloid load, tau load, and GFAP staining) and brain transcriptomic eigengenes for WGCNA-inferred modules (saddlebrown, mediumpurple3, orange, and darkgrey) using Hmisc::rcorr (version 4.4.2).

The major strength of this study is the paired biopsy data, and as such we did not approach our data from the standpoint of attempting to identify novel biomarkers. Instead, our goal was to identify proteins that have been well validated in prior work and examine how these markers correlate with our biopsy data. Using this approach, we started with a recently published analysis of AD CSF that also included data from four additional published validation datasets (5 cohorts total [20-22]). We focused on proteins in our own data that significantly increased or decreased in AD in at least one of the cohorts from this study with a corrected p-value (FDR) of 0.05, and at least nominally (unadjusted p-value less than 0.05) trended in a similar direction in at least one other cohort. This resulted in 45 proteins. We then tested whether any of these 45 proteins were nominally significantly correlated with either the histologic measurements of β-amyloid or tau on biopsy or any of our four gene expression modules. For module correlations, we only considered CSF proteins that were significantly correlated with a module eigengene if the same gene also correlated significantly and in a similar direction with the module eigengene in the brain RNA-seq data, as this would suggest that the CSF protein correlation is reflective of underlying CNS biology. Using these thresholds, we focused on 7 proteins from the proteomics data for further analysis. Note that all 7 CSF proteins that passed our criteria are correlating with our biopsy data in a similar direction to the reported correlations in the AD literature (i.e. if a CSF protein is reported to increase in AD, in our data it positively correlates with either AD pathology or a module that increases in AD, and vice versa). We did not apply an explicit threshold that this be the case in our filtering, and this supports the idea that our filtering has focused on CSF biomarkers of disease relevant for AD. One caveat to our proteomics analysis is that because of our filtering, we are not performing multiple testing correction (MTC) across all 607 proteins initially detected in our mass spectrometry data, and what we report in Figure 2A are unadjusted p-values. Many groups in the proteomics field have realized that the uncritical application of MTC for some datasets will result in a failure to detect any true positives even when many exist [23]. This has led several AD-related studies to use alternatives to MTC [24-27], and a recent meta-analysis indicated that useful data can be derived even in the face of limited statistical power for CSF proteomes [28]. Our confidence in our findings is supported in part by the filtering we have already done, and we are only focusing on the 7 proteins in our data that reliably trend similarly in at least two other AD studies.

**Figure 2.**
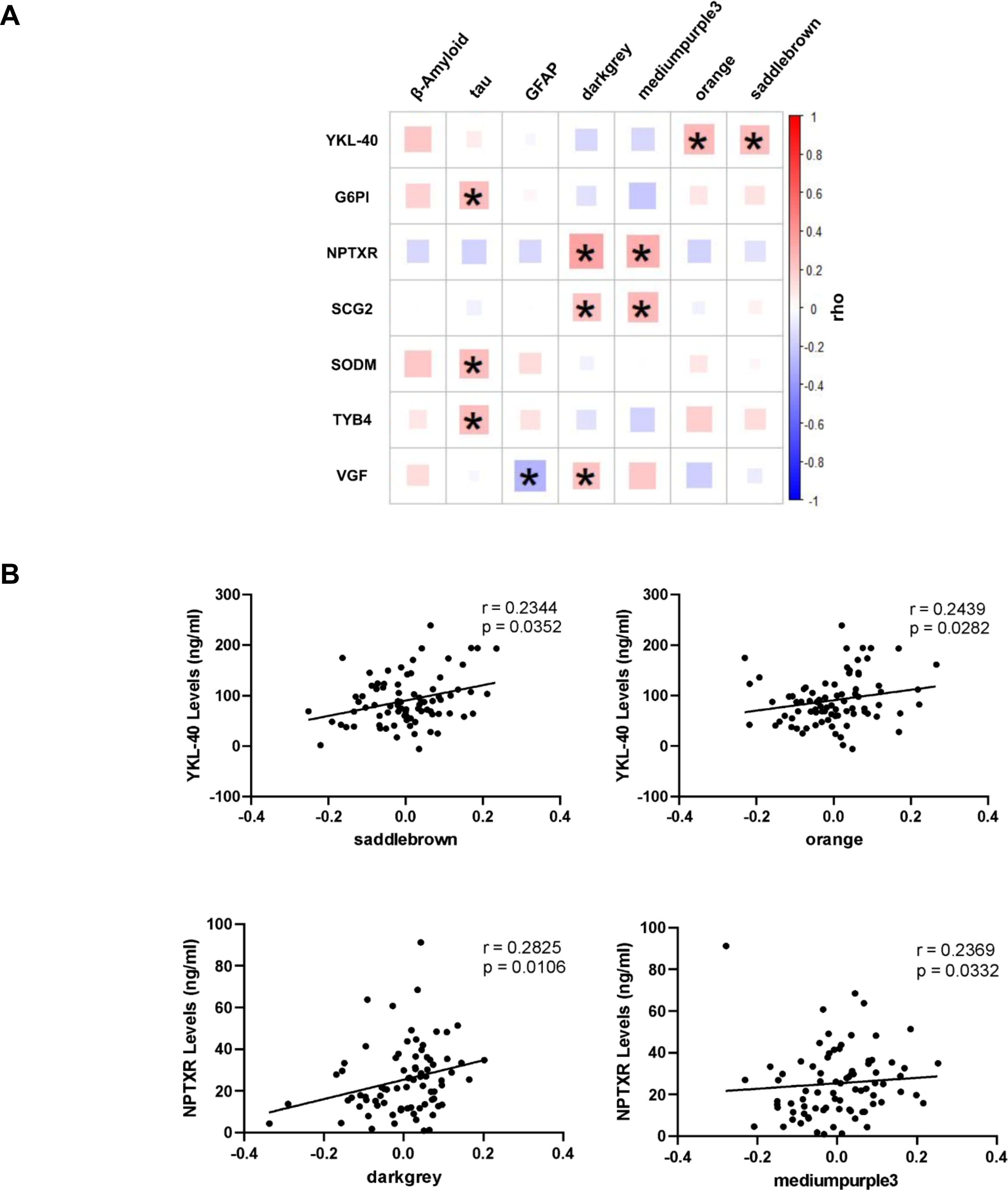
CSF proteomics data correlates with biopsy pathology and gene expression modules. Seven core proteins are shown that pass our filters for reproducibility (see Methods). In **A)** we show the Spearman’s correlations of these proteins with the eigengene of the four modules and quantified β-amyloid, tau, and GFAP on biopsy. * = p-values < 0.05. In **B)** we show Spearman’s correlation of 81 CSF samples with YKL-40 ELISA values vs. saddlebrown and orange module eigengenes and NPTXR ELISA values vs. darkgrey and mediumpurple3 module eigengenes. r and p values indicated. See Supplementary Tables 5 and 6 for numbers related to this figure.

In an effort to further reinforce our findings, we identified commercially available ELISAs that have been used extensively in the literature for two proteins of interest (YKL-40 and NPTXR) [29-34]. We performed ELISA measurements for these two proteins on CSF aliquots from all 81 subjects for ELISA validation, according to the manufacturers’ instructions. Levels of NPTXR (#ELH-NPTXR; Ray Biotech, GA, USA), and YKL-40 (# DC3L10; R&D systems, MN, USA), were assessed using a 1:10 and 1:100 dilution, respectively.

### 2.4 Metabolomics Analysis

The details on the acquisition of the untargeted metabolomics has been previously described [35, 36]. Briefly, the metabolites were extracted from plasma using acetonitrile and the extracts were injected in triplicate on two chromatographic columns: a hydrophilic interaction column (HILIC) under positive ionization (HILIC+) and a C18 column under negative ionization (C18-), to obtain three technical replicates per sample per column. Data was collected in full scan mode for molecules within 85-1250 kDa on a Thermo Orbitrap HFX Q-Exactive mass spectrometer. The untargeted metabolomic data were processed through a computational pipeline that leverages open source feature detection and peak alignment software, *apLCMS* [37] and *xMSanalyzer* [38]. Correction for batch effects was performed using ComBat, which uses an empirical Bayesian framework to adjust for known batches in which the samples were run [39]. Metabolic features detected in at least 70% of the samples were retained, leaving 3638 features from the HILIC+ column and 4532 features from the C18-column for further analysis. Zero-intensity values were considered below the detection limit of the instrument and were imputed with half the minimum intensity observed for each metabolic feature. The intensity of each metabolic feature was log10 transformed, quantile normalized, and auto-scaled for normalization and standardization. Normalized feature values for HILIC+ and C18-columns were regressed for sex and age and spearman correlated with each variable of interest (histological β-amyloid, tau, and GFAP, and the 4 modules). Correlation values of metabolic features with each variable of interest from each column were combined and fed into mummichog (MetaboAnalyst version 5.0) [40] to highlight significantly enriched metabolomic pathways (Gamma FDR adjusted p < 0.05). For the mummichog algorithm parameters, only pathways containing at least 5 metabolites were considered.

### 2.5 Statistics

All statistical analyses were performed in R (version 4.3.0), except those conducted with ELISA data which were performed using GraphPad Prism software (version 9.4.1). All correlations were performed using Spearman’s Rank Correlation Coefficient. Correlations for metabolomics and proteomics were assessed using the rcorr function as implemented in the Hmisc package (version 4.4.2) in R. FDR correction was used to adjust p-values for multiple comparisons where indicated. Hub genes were identified using the intramodularConnectivity function as implemented in WGCNA (version 1.72.1). The TAMPOR algorithm used for batch correction for the proteomics data can be downloaded from https://github.com/edammer/TAMPOR. Regressions for the proteomics and simoa data were performed using the function removeBatchEffect (version 3.54.2) as implemented in the package limma in R. Regressions for the metabolomics data were performed using the function ComBat as implemented in the package sva in R. Pathway enrichment analysis for the metabolomics data was conducted by the Mummichog algorithm in MetaboAnalyst (version 5.0). Of the 81 CSF samples, all analyses were performed successfully on the full set of 81 with the following exceptions; 78 have CSF Aβ40/42 values, 80 have CSF ptau 181 values, 77 have CSF ptau 181/Aβ42 values, 80 have CSF tau values, and 80 have immunohistochemical GFAP values.

### 2.6 Immunohistochemistry

Immunohistochemistry for GFAP was performed on sections of formalin fixed, paraffin embedded tissue using the rabbit monoclonal EP672Y antibody run on the Ventana Ultra platform. Brightfield microscopy was used to capture images of the GFAP stained slides at 4x magnification. Images were processed with CellProfiler v4.2.5 [41]. Tissue region within each image was distinguished from slide background through global minimum cross entropy thresholding, excluding objects below 50 pixels and any object that did not overlap with a manually drawn outline of the tissue region. Regions of histological artefact to exclude from tissue regions such as folded sections were identified through the global robust background thresholding for pixels over 2 standard deviations above the mean after discarding 85% of the bottom intensity pixels. Average GFAP intensity was then measured over the entire thresholded tissue area for each case. We were able to perform GFAP staining on 80 of the 81 specimens.

## 3. Results

### 3.1 Biopsy histologic changes and gene expression modules correlate with AD CSF biomarkers

We first sought to determine how pathology and transcriptomic changes in biopsy tissue relate to established AD biomarkers in CSF. In Figure 1B, we show that all four modules originally identified in [10] significantly correlate with previously quantified β-amyloid and tau pathology in our cohort of 81 subjects used in this study, similarly to the original group of 106 (WGCNA modules are labeled by color names, and we reproduce the same color names for these modules from [10]). In this study we have added quantified GFAP staining as a measure of astrogliosis in order to assess how this metric might also relate to our CSF analysis. Here, we note that GFAP staining correlates not only with the orange (astrocytic) module, but also with the darkgrey module, suggesting that neuronal function may decline in tandem with astrogliosis. To examine how our four modules and the quantified pathology on biopsy related to well-established AD CSF markers, we first performed Simoa measurements for β-amyloid-42 and - 40, total tau, phospho-tau-181 (p-tau181), and neurofilament light chain (NfL) (Figure 1C). Quantified β-amyloid plaques and tau pathology on biopsy correlate with Simoa measurements of β-amyloid-40/42 ratio, which is a superior biomarker for AD than β-amyloid-42 alone[42]. Tau pathology also correlates significantly with NfL. Both β-amyloid plaques and tau pathology trend positively with p-tau181 but are not significant, and p-tau181/β-amyloid-42 positively correlates with both. We next examined whether our modules correlated with any of the above Simoa measurements. All four modules have trends with β-amyloid-40/42, p-tau181, and p-tau181/β-amyloid-42 in directions consistent with the modules’ relationship with AD pathology, with modules that positively correlate with pathology having trends similar to β-amyloid plaque and tau pathology, and modules that decline with increasing pathology showing opposing trends. However, only correlations with NfL pass significance with three of the modules, with saddlebrown and orange positively correlating with NfL, and darkgrey negatively correlating with NfL.

In summary, all of the correlations found between the gene expression modules, pathology on biopsy, and Simoa metrics are internally consistent and support the view that the CSF of these patients is reflective of the ongoing AD-related disease captured on biopsy. Also encouragingly, this suggests that analysis of a very small piece of cortical tissue can give information that is partially predictive of CSF analysis. This should not immediately be assumed, as the CSF presumably captures changes found throughout the neuraxis, and may not correspond to the highly local analysis seen in a small piece of tissue. The logical reciprocal argument is that the findings in our biopsies are in fact reflecting similar biology seen in other areas of cortex, and that this is not only true of the measured pathology, but also of the gene expression data. Interestingly, GFAP staining does not appear to reach significance with any of the Simoa markers, although GFAP staining does correlate with quantified β-amyloid plaque pathology (Spearman’s r = 0.22, p-value = 0.047) and tau pathology (Spearman’s r = 0.32, p-value = 0.0044), suggesting that astrogliosis in these biopsies is at least partially reflective of AD pathology, particularly tau pathology.

### 3.2 CSF proteomics identifies concordant changes between AD-related CSF proteins and transcriptomic and pathologic changes in brain

We next performed label-free proteomics on our CSF samples. Our study is not as well powered as some prior analyses [20-22], and the major strength of this study is the paired biopsy data. Therefore, we did not approach our data from the standpoint of attempting to identify novel biomarkers, and instead used our data to study proteins that have been well validated in prior work and examine how these proteins correlate with our biopsy data. Specifically, we selected proteins that passed an FDR of 0.05 in at least one study and trended in the same direction (i.e. up or down in AD) with an unadjusted p-value of 0.05 in at least one other study, and which also correlated with one of our pathology variables or gene expression modules with an unadjusted p-value of 0.05. (see Methods for all details of our filtering steps). We focused on 7 proteins from the proteomics data that passed these thresholds for further analysis. In Figure 2A, we show these proteins and how they correlate with the biopsy data. Three neuronal proteins that have been shown to decrease in AD (NPTXR, SCG2, and VGF) [28], all positively correlate with darkgrey (our neuronal module), which is also consistent with darkgrey declining in tandem with AD pathology on biopsy (Figure 1; see Supplementary Table 2 for reproduced module genes). Mediumpurple3 (our microglial module enriched for homeostatic genes, which negatively correlates with AD pathology), also positively correlates with two of the neuronal markers, and GFAP negatively correlates with VGF. Tau pathology also positively correlates with several proteins previously shown to increase in AD CSF [20-22]. YKL-40 (otherwise known as CHI3L1) is a well-established marker of inflammation, and is widely studied in AD [28, 43]. In our data, this protein correlates with the orange (astrocytic) module and the saddlebrown (DAM microglial) module, suggesting that a multicellular response accompanies YKL-40 upregulation in CSF.

One caveat to our proteomics analysis is that we are not powered to perform multiple testing correction (MTC) across all proteins detected in our mass spectrometry data, and what we report in Figure 3 are unadjusted p-values. The variability of the CSF proteome is well documented [44], and it has been noted that the uncritical application of MTC for some datasets will result in a failure to detect any true positives even when many exist [23]. This has led several AD-related studies to use alternatives to MTC [24-27], and a recent meta-analysis indicated that useful data can be derived even in the face of limited statistical power for CSF proteomes [28]. Reproducibility across studies has been used as a filter in other analyses of AD CSF [28, 45], and here we have selected proteins that have reproducible trends across independent AD CSF studies and also significantly correlate with one of our AD-related variables. Note that all 7 CSF proteins that passed our criteria are correlating with our biopsy data in a similar direction to the reported correlations in the AD literature (i.e. if a CSF protein is reported to increase in AD, in our data it positively correlates with either AD pathology or a module that increases in AD, and vice versa). We did not apply an explicit threshold that this be the case in our filtering, and this supports the idea that our filtering has focused on CSF biomarkers of disease relevant for AD. Below, we also validate two key proteins with ELISAs that have been used extensively in the literature. Nevertheless, this lack of statistical power is a weakness of our study, which we further discuss in detail at the end of the Discussion section.

**Figure 3.**
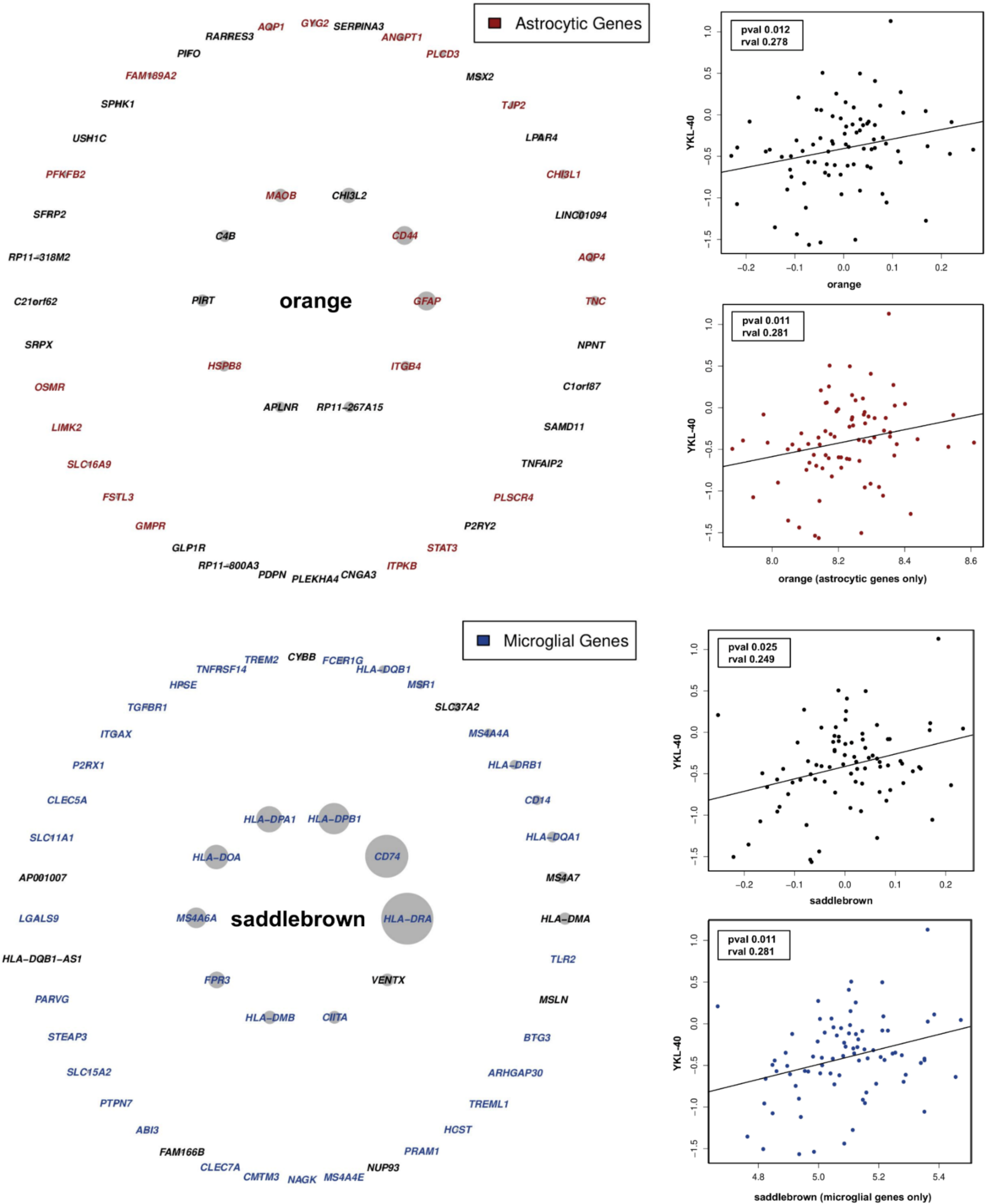
CSF YKL-40 correlates with microglial and astrocytic genes. Shown are the hub genes for the orange and saddlebrown modules, with astrocytic genes highlighted for orange and microglial genes highlighted for saddlebrown. Both modules correlate with CSF YKL-40. The mean gene expression vector of the astrocytic genes from orange and microglial genes from saddlebrown also correlate with YKL-40, supporting a role for these genes in the relationship between brain pathophysiology and CSF YKL-40. See Supplementary Table 7 for numbers used in this figure.

In an effort to further reinforce our findings, we identified commercially available ELISAs that have been used extensively in the literature for two proteins of interest (YKL-40 and NPTXR) [29-34]. We performed ELISA measurements for these two proteins on CSF aliquots from all 81 subjects in order to test the reliability of our proteomics data using a method orthogonal to mass spectrometry (in this case, sandwich ELISA). In Figure 2B, we show that ELISA measurements for these two proteins demonstrate a similar correlational profile to the proteomics data (additional correlations not shown are not significant - see Supplementary Table 6). Intriguingly, YKL-40 does not appear to correlate with AD pathology on biopsy. Although the lack of a significant correlation between CSF YKL-40 and AD pathology in biopsy tissue may be a power issue, at minimum it appears that in our cohort, CSF YKL-40 correlates more strongly with gene expression changes than AD pathology or traditional measures of astrogliosis (i.e. GFAP staining). As mentioned earlier, the saddlebrown and orange module are enriched for genes that are highly expressed in microglia and astrocytes respectively. In Figure 3, we show hub genes for each of these modules, with microglia and astrocytic genes highlighted in saddlebrown and orange respectively (hub genes are defined by intramodular connectivity [18], cell-type specific genes are from snRNA-seq data and are defined as enriched in a specific cell type compared to other cell types, which we have previously used to characterize these modules [10]). Mean gene expression vectors composed of only cell-type specific genes from these modules correlate as well with YKL-40 as the module eigengenes themselves, suggesting that the cell-type specific changes reported by these modules are correlating with YKL-40 (see Discussion).

### 3.3 CSF metabolomic analysis identifies pathways that are altered in the setting of early AD pathology

Finally, we analyzed our CSF with mass spectroscopy-based metabolomics to identify any biological pathways that may correspond to early AD pathology in brain tissue using mummichog, an analysis pipeline that identifies metabolic pathways enriched in metabolome correlations with variables of interest (see Methods). Pathways from several metabolic processes that have previously been reported in AD are also predicted to be altered in our CSF (Figure 4). Interestingly, fatty acid oxidation is predicted to be altered in tandem with β-amyloid pathology. Fatty acid oxidation has previously been linked to AD though several lines of investigation (reviewed in [46]). Fatty acid oxidation is relatively limited in neurons, in contrast to astrocytes, which metabolize fatty acids transported from neurons in ApoE-positive lipid particles as a protective mechanism in the setting of lipid peroxidation [47, 48]. Lipoic acid is an antioxidant and cofactor for several metabolic enzymes, and similarly to fatty acid oxidation, is linked to the generation of acetyl-CoA [49, 50], suggesting that there may be alterations in oxidative metabolism across multiple pathways as β-amyloid accumulates in the brain.

**Figure 4.**
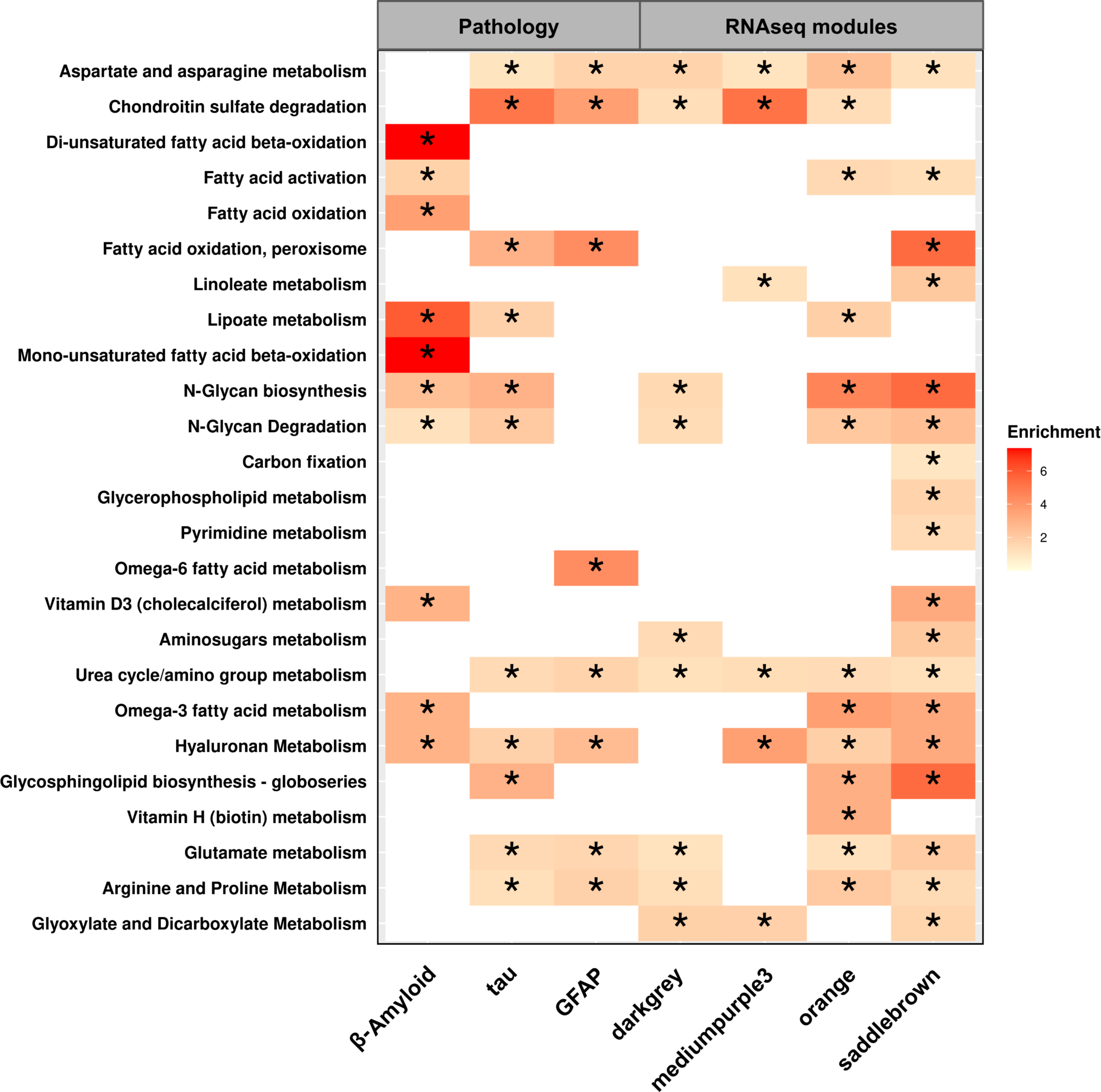
CSF metabolomics highlights AD relevant pathways. We analyzed our CSF with high-resolution mass spectroscopy-based metabolomics in order to identify any biological pathways that may correspond to early AD pathology in brain tissue using mummichog (see Methods). Representative pathways that are predicted to be altered in tandem with AD histology and gene expression changes are shown. * = FDR p-values < 0.05. See Supplementary Table 8 for numbers used in this figure.

Additional findings in our metabolomics data are alterations in aspartate and asparagine metabolism; marked alterations in aspartate metabolism have recently been found in AD brain tissue using metabolomics analysis [51]. N-glycan degradation is also significantly affected. Glycosylation abnormalities are common in AD, and a variety of abnormalities have been described [52, 53]. Tau undergoes glycosylation in AD brain tissue and not in control brain tissue, and this has been shown to be important for maintenance of paired helical filament structure [54]. In summary, several metabolic pathways that are relevant to AD are predicted to be altered in our CSF analysis, and suggest future avenues for investigating how these changes may correlate with AD pathophysiology (see Supplementary Table 8 for all pathways identified).

## 4. Discussion

The goal of this study is to address the major challenge of identifying biomarkers for established pathophysiologic CNS changes that occur in the setting of early AD pathology. Of particular interest is our finding that YKL-40 correlates primarily with two modules enriched for astrocytic and microglial genes. The interaction between microglia and astrocytes in AD is an area of active research, and recent work suggests that this interplay may contribute to neurodegeneration [55, 56]. YKL-40 is secreted by astrocytes, although its secretion is thought to be in part modulated through activated microglia [57, 58], which further supports our data showing that specific gene expression changes relating to these two cell types correlate with CSF YKL-40. Of note, our orange and saddlebrown modules are also highly correlated (r = 0.47, p-value = 5.066612E-07). One could speculate that we may be partially capturing a disease-relevant interplay between these two cell types in our data, and YKL-40 may be a useful marker to track this process. Future work could use these findings to examine this hypothesis in AD model systems. Our data also shows several CSF neuronal proteins that decrease in parallel with the darkgrey (neuronal) module. Interestingly, GFAP staining also (inversely) correlates with our neuronal module as well as VGF, and mediumpurple3 (our homeostatic microglial module) declines in parallel with some of the neuronal CSF proteins. This also points to aspects of the microglial/astrocytic response that may be most proximal to early neuronal dysfunction.

Although this study offers a unique opportunity to correlate analysis of CSF and brain tissue taken at the same timepoint from living patients, there are important limitations. First of all, unlike some large-scale studies [20, 22] we are not powered to perform MTC on our proteomics data. As mentioned earlier, this is a common problem with proteomics data from smaller cohorts, and other groups have reported unadjuted p-values and used alternate rationales for validation [23-28], including significant protein-protein interactions [24], coherent ontology groupings [27], and utility with predictive algorithms [25, 26]. Here, we have relied on prior validation in AD CSF in at least two other studies, as well as ELISA validation for two of our markers. We consider the two proteins validated by ELISA (NPTXR and YKL-40) to be the proteins where we are able to make the strongest argument concerning relationships with brain transcriptomic data. The other findings from our proteomics data are trends we consider reportable, but requiring additional validation in a future study. To our knowledge, this is the first study to directly link CSF biomarkers and AD-related changes in CNS gene expression in the same patients. While this certainly lends novelty to our findings, it also limits us in options to validate our results. As NPH cohorts are increasingly studied by AD researchers, there will hopefully be more reports where cross-comparision with the data presented here is possible.

In addition, all of the patients in this study have the comorbitidy of hydrocephalus. Although it is not easy to disentangle what effect this might have on the data presented here, it should be noted that AD is usually accompanied by co-morbid neurologic disease, and that “pure AD” actually constitutes a minority of AD cases [59-61]. Thus, pure AD is actually less common among patients with dementia than mixed pathology, and there is no *a priori* reason to expect hydrocephalus to uniquely affect our analysis more than other common confounders found in various autopsy and clinical cohorts. Although the interaction of NPH and AD is an area of ongoing research [5, 8, 9], the fact that we find correlations between AD pathology on biopsy and AD biomarkers in CSF taken at the same timepoint is itself internal validation that aspects of AD pathophysiology can be studied using our approach, even in the presence of potential confounders.

In summary, we show for the first time how CNS transcriptomic changes (and accompanying early AD pathology) are related to CSF biomarkers. As new disease-modifying therapies are developed targeting specific physiologic aspects of AD (such as synaptic dysfunction or the immune response), biomarkers that track these changes will be crucial. The data presented here offers both biomarkers that can be used for these purposes as well directions for future work.

## Data Availability

All data produced in the present study is available upon reasonable request to the authors.

## Abbreviations

AD: Alzheimer’s disease
NPH: Normal pressure hydrocephalus

## Consent Statement

This study is a retrospective study that uses tissue and CSF samples not required for clinical diagnosis and associated clinical and demographic data. Patients undergoing surgery signed an umbrella consent form, providing consent to use the remaining biopsies and body fluids for research. This study was reviewed and approved by the Columbia University Institutional Review Board (IRB), and all relevant ethical regulations have been followed.

---

**Supplementary Figure 1.**
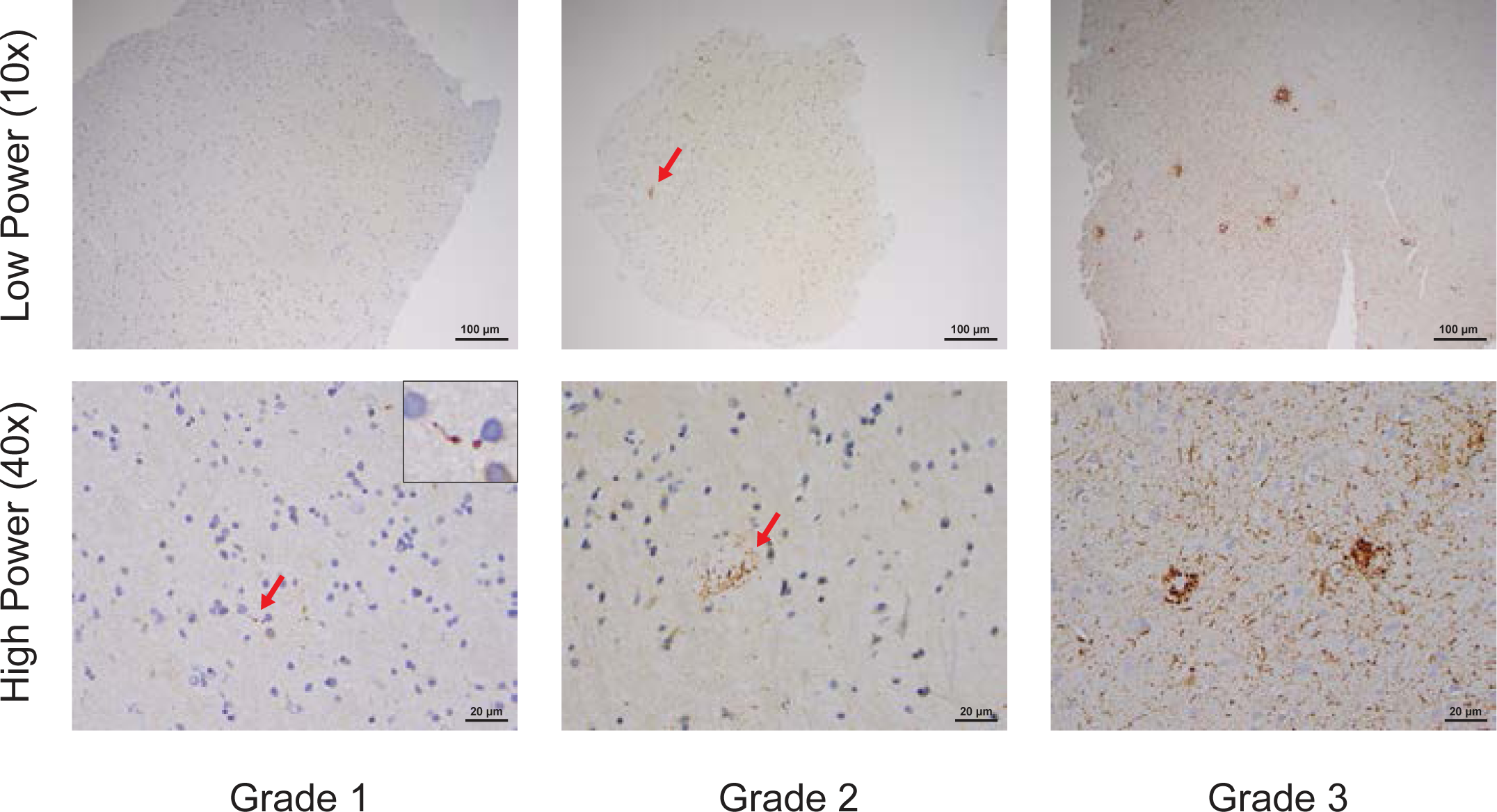
The biopsies in our cohort have minimal tau pathology on immunohistochemistry. (reproduced from [10]) - We devised a rating scale to grade the minimal degree of tau pathology seen in NPH biopsies. All panels display tau immunohistochemistry of NPH biopsies at low-power (10x magnification, upper row) or high-power (40x magnification, lower row). Grade 0 was given to biopsies with no tau pathology. Grade 1 (left panels) was given to biopsies that have any tau pathology at all, usually one or more dystrophic neurites, but do not make criteria for Grade 2 (middle panels). Grade 2 was given to biopsies that have at least one tau-positive neuron or neuritic plaque, but do not make criteria for Grade 3 (right panels). Grade 3 was reserved for biopsies with tau pathology evenly distributed throughout the biopsy. It is difficult to appreciate tau pathology at low-power in grade 1 slides (upper left panel), and high-power is required to see single dystrophic neurites (lower left panel - red arrow, see blow-up insert). In grade 2 slides, one can usually see neuritic plaques at low-power (middle upper panel, red arrow), as well as at high-power (middle lower panel, red arrow). Grade 3 slides have obvious tau pathology at all magnifications.

## References

[1] Oliveira LM, Nitrini R, Roman GC. Normal-pressure hydrocephalus: A critical review. Dement Neuropsychol. 2019;13:133–43.

[2] Williams MA, Malm J. Diagnosis and Treatment of Idiopathic Normal Pressure Hydrocephalus. Continuum (Minneap Minn). 2016;22:579–99.

[3] Borgesen SE, Gjerris F. The predictive value of conductance to outflow of CSF in normal pressure hydrocephalus. Brain. 1982;105:65–86.

[4] Symon L, Dorsch NW. Use of long-term intracranial pressure measurement to assess hydrocephalic patients prior to shunt surgery. Journal of neurosurgery. 1975;42:258–73.

[5] McGovern RA, Nelp TB, Kelly KM, Chan AK, Mazzoni P, Sheth SA, et al. Predicting Cognitive Improvement in Normal Pressure Hydrocephalus Patients Using Preoperative Neuropsychological Testing and Cerebrospinal Fluid Biomarkers. Neurosurgery. 2019;85:E662–E9.

[6] Hamilton R, Patel S, Lee EB, Jackson EM, Lopinto J, Arnold SE, et al. Lack of shunt response in suspected idiopathic normal pressure hydrocephalus with Alzheimer disease pathology. Ann Neurol. 2010;68:535–40.

[7] Libard S, Alafuzoff I. Alzheimer’s disease neuropathological change and loss of matrix/neuropil in patients with idiopathic Normal Pressure Hydrocephalus, a model of Alzheimer’s disease. Acta neuropathologica communications. 2019;7:98.

[8] Leinonen V, Koivisto AM, Savolainen S, Rummukainen J, Tamminen JN, Tillgren T, et al. Amyloid and tau proteins in cortical brain biopsy and Alzheimer’s disease. Ann Neurol. 2010;68:446–53.

[9] Luikku AJ, Hall A, Nerg O, Koivisto AM, Hiltunen M, Helisalmi S, et al. Predicting Development of Alzheimer’s Disease in Patients with Shunted Idiopathic Normal Pressure Hydrocephalus. J Alzheimers Dis. 2019;71:1233–43.

[10] Huang W, Bartosch AM, Xiao H, Maji S, Youth EHH, Flowers X, et al. An immune response characterizes early Alzheimer’s disease pathology and subjective cognitive impairment in hydrocephalus biopsies. Nat Commun. 2021;12:5659.

[11] Gazestani V, Kamath T, Nadaf NM, Dougalis A, Burris SJ, Rooney B, et al. Early Alzheimer’s disease pathology in human cortex involves transient cell states. Cell. 2023;186:4438–53 e23.

[12] Keren-Shaul H, Spinrad A, Weiner A, Matcovitch-Natan O, Dvir-Szternfeld R, Ulland TK, et al. A Unique Microglia Type Associated with Restricting Development of Alzheimer’s Disease. Cell. 2017;169:1276–90 e17.

[13] Gerrits E, Brouwer N, Kooistra SM, Woodbury ME, Vermeiren Y, Lambourne M, et al. Distinct amyloid-beta and tau-associated microglia profiles in Alzheimer’s disease. Acta Neuropathol. 2021;141:681–96.

[14] S. A. FastQC: a quality control tool for high throughput sequence data.

[15] Durbin BP, Hardin JS, Hawkins DM, Rocke DM. A variance-stabilizing transformation for gene-expression microarray data. Bioinformatics. 2002;18 Suppl 1:S105–10.

[16] Leek JT, Storey JD. Capturing heterogeneity in gene expression studies by surrogate variable analysis. PLoS Genet. 2007;3:1724–35.

[17] Ritchie ME, Phipson B, Wu D, Hu Y, Law CW, Shi W, et al. limma powers differential expression analyses for RNA-sequencing and microarray studies. Nucleic Acids Res. 2015;43:e47.

[18] Langfelder P, Horvath S. WGCNA: an R package for weighted correlation network analysis. BMC Bioinformatics. 2008;9:559.

[19] Szeto B, Valentini C, Aksit A, Werth EG, Goeta S, Brown LM, et al. Impact of Systemic versus Intratympanic Dexamethasone Administration on the Perilymph Proteome. J Proteome Res. 2021;20:4001–9.

[20] Johnson ECB, Dammer EB, Duong DM, Ping L, Zhou M, Yin L, et al. Large-scale proteomic analysis of Alzheimer’s disease brain and cerebrospinal fluid reveals early changes in energy metabolism associated with microglia and astrocyte activation. Nature medicine. 2020;26:769–80.

[21] Dayon L, Nunez Galindo A, Wojcik J, Cominetti O, Corthesy J, Oikonomidi A, et al. Alzheimer disease pathology and the cerebrospinal fluid proteome. Alzheimers Res Ther. 2018;10:66.

[22] Higginbotham L, Ping L, Dammer EB, Duong DM, Zhou M, Gearing M, et al. Integrated proteomics reveals brain-based cerebrospinal fluid biomarkers in asymptomatic and symptomatic Alzheimer’s disease. Sci Adv. 2020;6.

[23] Pascovici D, Handler DC, Wu JX, Haynes PA. Multiple testing corrections in quantitative proteomics: A useful but blunt tool. Proteomics. 2016;16:2448–53.

[24] Loupy KM, Lee T, Zambrano CA, Elsayed AI, D’Angelo HM, Fonken LK, et al. Alzheimer’s Disease: Protective Effects of Mycobacterium vaccae, a Soil-Derived Mycobacterium with Anti-Inflammatory and Anti-Tubercular Properties, on the Proteomic Profiles of Plasma and Cerebrospinal Fluid in Rats. J Alzheimers Dis. 2020;78:965–87.

[25] Khan MJ, Desaire H, Lopez OL, Kamboh MI, Robinson RAS. Why Inclusion Matters for Alzheimer’s Disease Biomarker Discovery in Plasma. J Alzheimers Dis. 2021;79:1327–44.

[26] Kim Y, Kim J, Son M, Lee J, Yeo I, Choi KY, et al. Plasma protein biomarker model for screening Alzheimer disease using multiple reaction monitoring-mass spectrometry. Scientific reports. 2022;12:1282.

[27] King CD, Robinson RAS. Evaluating Combined Precursor Isotopic Labeling and Isobaric Tagging Performance on Orbitraps To Study the Peripheral Proteome of Alzheimer’s Disease. Anal Chem. 2020;92:2911–6.

[28] Pedrero-Prieto CM, Garcia-Carpintero S, Frontinan-Rubio J, Llanos-Gonzalez E, Aguilera Garcia C, Alcain FJ, et al. A comprehensive systematic review of CSF proteins and peptides that define Alzheimer’s disease. Clin Proteomics. 2020;17:21.

[29] Babic Leko M, Mihelcic M, Jurasovic J, Nikolac Perkovic M, Spanic E, Sekovanic A, et al. Heavy Metals and Essential Metals Are Associated with Cerebrospinal Fluid Biomarkers of Alzheimer’s Disease. Int J Mol Sci. 2022;24.

[30] Dulewicz M, Kulczynska-Przybik A, Borawska R, Slowik A, Mroczko B. Evaluation of Synaptic and Axonal Dysfunction Biomarkers in Alzheimer’s Disease and Mild Cognitive Impairment Based on CSF and Bioinformatic Analysis. Int J Mol Sci. 2022;23.

[31] Kusnierova P, Zeman D, Hradilek P, Zapletalova O, Stejskal D. Determination of chitinase 3-like 1 in cerebrospinal fluid in multiple sclerosis and other neurological diseases. PLoS One. 2020;15:e0233519.

[32] Lim B, Sando SB, Grontvedt GR, Brathen G, Diamandis EP. Cerebrospinal fluid neuronal pentraxin receptor as a biomarker of long-term progression of Alzheimer’s disease: a 24-month follow-up study. Neurobiol Aging. 2020;93:97 e1–e7.

[33] Teitsdottir UD, Jonsdottir MK, Lund SH, Darreh-Shori T, Snaedal J, Petersen PH. Association of glial and neuronal degeneration markers with Alzheimer’s disease cerebrospinal fluid profile and cognitive functions. Alzheimers Res Ther. 2020;12:92.

[34] Dulewicz M, Kulczynska-Przybik A, Slowik A, Borawska R, Mroczko B. Neurogranin and Neuronal Pentraxin Receptor as Synaptic Dysfunction Biomarkers in Alzheimer’s Disease. J Clin Med. 2021;10.

[35] Liu KH, Walker DI, Uppal K, Tran V, Rohrbeck P, Mallon TM, et al. High-Resolution Metabolomics Assessment of Military Personnel: Evaluating Analytical Strategies for Chemical Detection. J Occup Environ Med. 2016;58:S53–61.

[36] Vardarajan B, Kalia V, Manly J, Brickman A, Reyes-Dumeyer D, Lantigua R, et al. Differences in plasma metabolites related to Alzheimer’s disease, APOE epsilon4 status, and ethnicity. Alzheimers Dement (N Y). 2020;6:e12025.

[37] Yu T, Park Y, Johnson JM, Jones DP. apLCMS--adaptive processing of high-resolution LC/MS data. Bioinformatics. 2009;25:1930–6.

[38] Uppal K, Soltow QA, Strobel FH, Pittard WS, Gernert KM, Yu T, et al. xMSanalyzer: automated pipeline for improved feature detection and downstream analysis of large-scale, non-targeted metabolomics data. BMC Bioinformatics. 2013;14:15.

[39] Leek JT, Johnson WE, Parker HS, Jaffe AE, Storey JD. The sva package for removing batch effects and other unwanted variation in high-throughput experiments. Bioinformatics. 2012;28:882–3.

[40] Li S, Park Y, Duraisingham S, Strobel FH, Khan N, Soltow QA, et al. Predicting network activity from high throughput metabolomics. PLoS Comput Biol. 2013;9:e1003123.

[41] Stirling DR, Swain-Bowden MJ, Lucas AM, Carpenter AE, Cimini BA, Goodman A. CellProfiler 4: improvements in speed, utility and usability. BMC Bioinformatics. 2021;22:433.

[42] Hansson O, Lehmann S, Otto M, Zetterberg H, Lewczuk P. Advantages and disadvantages of the use of the CSF Amyloid beta (Abeta) 42/40 ratio in the diagnosis of Alzheimer’s Disease. Alzheimers Res Ther. 2019;11:34.

[43] Whelan CD, Mattsson N, Nagle MW, Vijayaraghavan S, Hyde C, Janelidze S, et al. Multiplex proteomics identifies novel CSF and plasma biomarkers of early Alzheimer’s disease. Acta neuropathologica communications. 2019;7:169.

[44] Schilde LM, Kosters S, Steinbach S, Schork K, Eisenacher M, Galozzi S, et al. Protein variability in cerebrospinal fluid and its possible implications for neurological protein biomarker research. PLoS One. 2018;13:e0206478.

[45] Wesenhagen KEJ, Teunissen CE, Visser PJ, Tijms BM. Cerebrospinal fluid proteomics and biological heterogeneity in Alzheimer’s disease: A literature review. Crit Rev Clin Lab Sci. 2020;57:86–98.

[46] Yin F. Lipid metabolism and Alzheimer’s disease: clinical evidence, mechanistic link and therapeutic promise. FEBS J. 2023;290:1420–53.

[47] Ioannou MS, Jackson J, Sheu SH, Chang CL, Weigel AV, Liu H, et al. Neuron-Astrocyte Metabolic Coupling Protects against Activity-Induced Fatty Acid Toxicity. Cell. 2019;177:1522–35 e14.

[48] Baxter PS, Hardingham GE. Adaptive regulation of the brain’s antioxidant defences by neurons and astrocytes. Free Radic Biol Med. 2016;100:147–52.

[49] Kohlmeier M. Nutrient metabolism. Amsterdam; Boston: Academic Press; 2003.

[50] Solmonson A, DeBerardinis RJ. Lipoic acid metabolism and mitochondrial redox regulation. The Journal of biological chemistry. 2018;293:7522–30.

[51] Paglia G, Stocchero M, Cacciatore S, Lai S, Angel P, Alam MT, et al. Unbiased Metabolomic Investigation of Alzheimer’s Disease Brain Points to Dysregulation of Mitochondrial Aspartate Metabolism. J Proteome Res. 2016;15:608–18.

[52] Haukedal H, Freude KK. Implications of Glycosylation in Alzheimer’s Disease. Front Neurosci. 2020;14:625348.

[53] Kizuka Y, Kitazume S, Taniguchi N. N-glycan and Alzheimer’s disease. Biochim Biophys Acta Gen Subj. 2017;1861:2447–54.

[54] Wang JZ, Grundke-Iqbal I, Iqbal K. Glycosylation of microtubule-associated protein tau: an abnormal posttranslational modification in Alzheimer’s disease. Nature medicine. 1996;2:871–5.

[55] Liddelow SA, Barres BA. Reactive Astrocytes: Production, Function, and Therapeutic Potential. Immunity. 2017;46:957–67.

[56] Liddelow SA, Guttenplan KA, Clarke LE, Bennett FC, Bohlen CJ, Schirmer L, et al. Neurotoxic reactive astrocytes are induced by activated microglia. Nature. 2017;541:481–7.

[57] Bonneh-Barkay D, Bissel SJ, Kofler J, Starkey A, Wang G, Wiley CA. Astrocyte and macrophage regulation of YKL-40 expression and cellular response in neuroinflammation. Brain Pathol. 2012;22:530–46.

[58] Connolly K, Lehoux M, O’Rourke R, Assetta B, Erdemir GA, Elias JA, et al. Potential role of chitinase-3-like protein 1 (CHI3L1/YKL-40) in neurodegeneration and Alzheimer’s disease. Alzheimers Dement. 2023;19:9–24.

[59] Schneider JA, Arvanitakis Z, Bang W, Bennett DA. Mixed brain pathologies account for most dementia cases in community-dwelling older persons. Neurology. 2007;69:2197–204.

[60] James BD, Wilson RS, Boyle PA, Trojanowski JQ, Bennett DA, Schneider JA. TDP-43 stage, mixed pathologies, and clinical Alzheimer’s-type dementia. Brain. 2016;139:2983–93.

[61] Barnes LL, Leurgans S, Aggarwal NT, Shah RC, Arvanitakis Z, James BD, et al. Mixed pathology is more likely in black than white decedents with Alzheimer dementia. Neurology. 2015;85:528–34.

